# Vascular Aging and Bladder Health: Associations Between Peripheral Arterial Disease and Lower Urinary Tract Symptoms in Mid-Life Women from CARDIA-PLUS

**DOI:** 10.64898/2026.10.02.26364598

**Authors:** Alayne D. Markland, Whitney K. Hendrickson, Andres Arguedas, Simion De, Katelyn Adams, C. Elizabeth Lewis, Gerhard Hellemann, Cynthia S. Fok, Sonya S. Brady

**Author notes:** **Corresponding author: Dr. Alayne Markland** University of Utah Department of Internal Medicine, Division of Geriatrics, 30 N Mario Capecchi Dr., Salt Lake City, UT.

## Abstract

**Objectives:** To describe associations between peripheral arterial disease (PAD) and bladder health (BH) and lower urinary tract symptoms (LUTS) among a subset of women in the Coronary Artery Risk Development In Young Adults (CARDIA) study.

**Methods:** Women from CARDIA at Birmingham, AL and Minneapolis, MN were eligible if they completed a questionnaire about bladder symptoms in 2022-23, an ancillary study post-year 35 called CARDIA-PLUS, including the global BH scale (possible range, 0-100) and the LURN-SI 10 (0-38) and the AUASI (0-35). A subset (n=216) completed an inperson examination, including the ankle brachial index (ABI) procedure (n=194). ABI scores were clinically categorized as normal (1.00-1.40), borderline abnormal (0.91-0.99), and abnormal (≤0.90; evidence of PAD). Continuous BH and LUTS variables were regressed on ABI category, adjusting for sociodemographics, obstetric/gynecologic variables, co-morbidities, and physical activity.

**Key Results:** Among women who completed BH/LUTS questionnaires and the ABI (n=192), the mean age was 63±4 years and racial composition was 47% Black and 53% White. ABI results were normal (74%), borderline abnormal (20%), and abnormal (6%). After adjusting for covariates, women with abnormal ABI scores had higher LURN-SI scores (4.1 points, 95% CI 0.7, 7.5) and did not have higher AUASI scores (1.4 points, 95% CI −3.1, 5.7) or lower BH scores (−9.6 points, 95% CI −24.0, 4.9) compared to women with normal ABI scores. Borderline abnormal ABI was not associated with BH or LUTS.

**Discussion/Conclusion:** In this cohort, PAD was associated with greater LUTS severity. PAD and LUTS may share common etiologies and risk factors that need additional research.

## Introduction

Bothersome lower urinary tract symptoms (LUTS), including overactive bladder (OAB) and urinary incontinence (UI) are common and increase with advancing age. Prevalence data indicate that more than half of adults experience LUTS at any given time—with prevalence rates ranging from 55% to 79% in women, depending on the population and criteria used.^1–4^ As the population of older adults in the U.S. increases, health care costs associated with LUTS and OAB continue to grow.^5^

There is strong and increasing evidence that cardiovascular (CV) risk may be associated with LUTS and support vascular aging mechanisms interconnecting macro-and micro-vascular changes.^6–12^ The existing literature on CV risk, cardiovascular disease (CVD), and LUTS is primarily in men, with significantly less data in women.^6–11^ In animal studies, macrovascular changes with induced atherosclerosis of the iliac arteries leads to decreased bladder perfusion at the microvascular level; depending on the severity of ischemia, detrusor overactivity or underactivity develops, both hallmarks of LUTS.^13–20^ Small pilot studies have demonstrated decreased perfusion to the bladder at the microvascular level, in those with LUTS.^21–25^ Therefore, CVD, specifically peripheral arterial disease (PAD), including pelvic vasculature, may be associated with LUTS through a shared mechanism of poor peripheral perfusion and pelvic ischemia. If associated, CV prevention strategies could inform prevention of LUTS, as well as PAD.^26,27^

To further research on CVD and LUTS to inform vascular geroscience, we evaluated PAD using the Ankle Brachial Index (ABI), a macrovascular measure of vascular aging, among a subset of mid-life women in an ancillary study to the Coronary Artery Risk Development In Young Adults (CARDIA) longitudinal cohort, called CARDIA-PLUS. Clinically, PAD is associated with coronary and cerebrovascular morbidity and measured in routine clinical care by ABI.^28^ Our primary objective was to describe associations of PAD with perceptions of bladder health (BH) and self-report of LUTS while also adjusting for key covariates. We hypothesized that mid-life women with PAD, defined by ABI score, would have more LUTS and lower rates of bladder health, and that these associations would be attenuated by adjustment for CVD risk factors.

## METHODS

Coronary Artery Risk Development In Young Adults (CARDIA) is a longitudinal cohort study that began in 1985-86 to examine the development of cardiovascular disease across the lifespan. The Year 35 assessment, 35 years post-baseline, was completed in 2020-2022. BH and LUTS questionnaires were mailed or electronically sent to participants from Birmingham, AL and Minneapolis, MN CARDIA field centers after Year 35 (2022-24) if they had completed prior LUTS questionnaires from 2012-2013 and completed the first phase of the Year 35 assessment. The University of Alabama at Birmingham provided single IRB approval for both sites, and all participants completed informed consent for this ancillary study.

### Recruitment for CARDIA-PLUS Clinical Examination after the Year 35 assessment

Women indicated interest in an in-person clinical examination after completion of the LUTS questionnaires. Interested women were scheduled for an in-person examination within 12 weeks of questionnaire completion. The in-person examination included completion of the ABI and other procedures. The analytic sample excluded women with prior lymphadenectomies, shunts, or grafts; women who experienced pain during the ABI; and women with non-compressible ABI values.

Figure 1 displays a flow chart of participants who were eligible for the present study and progressed to the analytic data set (i.e., had complete ABI and outcome data).

**FIGURE 1.**
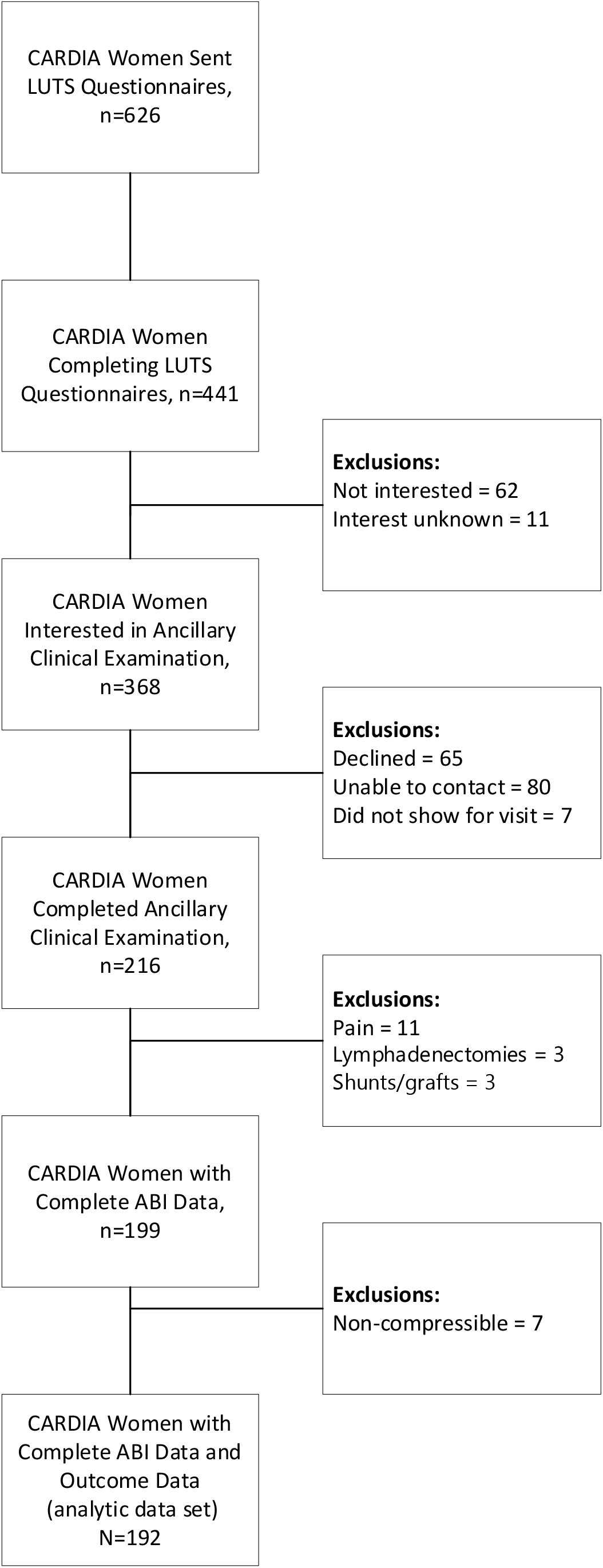
CARDIA Women Eligible for Clinical Examination and Exclusions

### ABI Examination and Scoring

The ABI procedure measures systolic blood pressure (SBP) in the brachial arteries of both arms, the dorsalis pedis (DP) artery of each leg, and the posterior tibial (PT) artery of each leg.^28^ Clinical staff utilized a standardized protocol to measure ABI and received centralized training prior to data collection.^28^ Women’s arms and legs were measured to ensure accurate cuff sizing for blood pressure (BP) measurements.

Women remained supine in a quiet examination room for approximately five minutes prior to beginning. A handheld Doppler instrument and a manual sphygmomanometer were used to obtain the SBP at each location in the following order: right brachial artery, right DP artery, right PT artery, left PT artery, left DP artery, left brachial artery. Clinical guidelines were used to define ABI classifications, where normal was defined as 1.00-1.40, borderline abnormal as 0.91-0.99 and abnormal as < 0.90, >1.40 defined noncompressible.^28^

### Bladder Health and LUTS

Bladder health (BH) was assessed using the RISE FOR HEALTH global bladder health subscale (6 items; e.g., How would you rate the function of your bladder?).^29^ Scores ranged from 0-100 with greater scores representing better perceived bladder health. The Lower Urinary Tract Research Network short inventory (LURN-SI) with 10 questions assessed the presence of LUTS, including UI and bladder pain.^30^ Scores ranged from 0-38 with higher scores representing a higher number of and more frequent LUTS. Presence and severity of other LUTS were assessed with the American Urologic Association Symptom Index (AUA-SI) score, a composite of 7 individual LUTS, with response options related to frequency: “not at all, less than 1 in 5 times, less than half the time, about half the time, more than half the time, and almost always” with scores ranging from 0-35 and higher scores representing higher severity.^31,32^

### Descriptive Variables and Covariates

Covariates were obtained from Year 35 CARDIA data, including sociodemographic variables (age, race, and educational attainment), obstetric and gynecologic history (parity, hysterectomy, and menopausal status), and medical history (hypertension from BP measurements or medication usage, diabetes from blood glucose or medication usage, and nonfatal coronary artery disease from adjudicated records). From the physical examination, participants had weight (kg), height (m), and waist circumference (cm) measured. Body mass index (kg/m^2^) was defined as normal <25, overweight ≥25-<30, and obesity if ≥30.0 kg/m^2^. As a descriptive variable, Cardiovascular-Kidney-Metabolic (CKM) syndrome is defined by the co-occurrence or progression of metabolic risk factors (visceral adiposity, dyslipidemia, prediabetes/hypertension), established metabolic disease or CKD, subclinical cardiovascular disease, and ultimately clinical CVD. Women with normal BMI and waist circumference (<88 cm), normoglycemia (fasting blood glucose <100 mg/dL or HbA1c between <5.7%, normotension (systolic blood pressure <130 mm Hg or diastolic blood pressure <80 mm Hg and without use of antihypertensive medications), a normal lipid profile (HDL ≥50 mg/dL; hypertriglyceridemia <135 mg/dL, and no evidence of chronic kidney disease or subclinical or clinical CVD were compared to women who met criteria for CKM. Physical activity was assessed from the CARDIA Physical Activity History questionnaire. This questionnaire assessed activity in 13 specific activities over the previous year, including sports, exercise, home maintenance, and occupational activities, classifying each as nonparticipation, infrequent, or frequent based on monthly hours of engagement. Responses are converted into exercise units derived by multiplying activity intensity (in MET-minutes) by weighted months of infrequent and frequent participation. ^33^

### Statistical Analysis

Of the 662 CARDIA women in AL and MN, 441 women from AL and MN enrolled in CARDIA-PLUS, 216 completed the clinical examination and 199 had ABI data with 192 in our analytic sample (Figure 1) per STROBE guidelines.^34^ We compared women’s descriptive statistics across the ABI groups using Pearson’s Chi-square test, Kruskal-Wallis tests and Fisher’s exact test for p values. We then constructed separate linear regression models of each BH and LUTS variable on ABI classification, adjusting for covariates in stages. ABI score was examined as a categorical variable with three categories (normal, borderline abnormal, and abnormal); women with normal scores served as the reference group in regression analyses.

## RESULTS

Of the 192 women, mean age was 63.6 (standard deviation [SD] 3.5) years old with 47% Black women and 53% White women (Table 1). Half had hypertension (50%), 46% had a BMI > 30 kg/m^2^, and 21% had diabetes mellitus.

**TABLE 1.** Descriptive Data for Women by ABI Classification Groups.

|  | Total | ABI Classifications, n (%) |  |  |  |
| --- | --- | --- | --- | --- | --- |
| Variable | N = 192 | Normal, 1.00-1.40, n=142 (74) | Borderline, 0.91-0.99, n=38 (20) | Abnormal, ≤0.90, n=12 (6) | p-value<sup>1</sup> |
| **Sociodemographics** |  |  |  |  |  |
| Age in years, mean (SD) | 63.6 (3.5) | 63.46 (3.6) | 63.8 (3.2) | 64.2 (3.1) | 0.80 |
| Race, n (%) |  |  |  |  | 0.91 |
| Black | 91 (47) | 66 (46) | 19 (50) | 6 (50) |  |
| White | 101 (53) | 76 (54) | 19 (50) | 6 (50) |  |
| Education, n (%) |  |  |  |  | 0.75 |
| College graduate | 95 (50) | 71 (50) | 19 (50) | 5 (42) |  |
| High School or less | 34 (18) | 23 (16) | 9 (24) | 2 (16) |  |
| Some college | 63 (32) | 48 (34) | 10 (26) | 5 (42) |  |
| **Obstetric and Gynecologic Factors** |  |  |  |  |  |
| Parity, mean (SD) | 1.6 (1.3) | 1.7 (1.3) | 1.5 (1.3) | 1.6 (0.9) | 0.89 |
| Hysterectomy, n (%) | 65 (35) | 49 (36) | 10 (27) | 6 (55) | 0.24 |
| Menopause, n (%) | 82 (43) | 60 (43) | 17 (46) | 5 (46) | 0.97 |
| Not sure | 33 (17) | 26 (18) | 5 (14) | 2 (18) |  |
| **Behavioral and Medical Comorbidities** |  |  |  |  |  |
| BMI, mean (SD) | 30.5 (7.1) | 30.4 (6.9) | 30.8 (8.25) | 31.8 (4.8) | 0.52 |
| Obesity, n (%) | 88 (46) | 62 (44) | 18 (47) | 8 (67) | 0.30 |
| Waist circumference (cm),<br>mean (SD) | 92.3 (14.3) | 91.7 (14.1) | 92.8 (15.6) | 97.4 (12.4) | 0.35 |
| HTN, n (%) | 91 (50) | 67 (49) | 16 (44) | 8 (80) | 0.13 |
| DM, n (%) | 40 (21) | 39 (20) | 5 (13) | 6 (50) | 0.03 |
| CAD, nonfatal, n (%) | 10 (5) | 6 (4) | 3 (8) | 1 (8) | 0.39 |
| Cardio-kidney Metabolic<br>syndrome, n (%) | 51 (27) | 35 (25) | 12 (32) | 4 (33) | 0.57 |
| Physical activity, mean (SD)<br>in Exercise Units | 307.5 (253.0) | 319.2 (261.7) | 261.0 (207.8) | 316.9 (281.5) | 0.60 |
| **Bladder Health and LUTS Scores** |  |  |  |  |  |
| Global Bladder Health, mean<br>(SD), scale 0-100, higher<br>scores representing better<br>health | 68.0 (20.3) | 68.9 (19.7) | 67.7 (20.3) | 58.7 (26.8) | 0.45 |
| LURN-10 scores, mean (SD),<br>scale 0-38, higher scores<br>representing more<br>symptoms | 7.2 (5.0) | 7.0 (4.5) | 7.1 (4.4) | 11.4 (9.5) | 0.52 |
| AUASI Scores, mean (SD),<br>scale 0-35, higher scores<br>representing more<br>symptoms | 9.0 (6.2) | 8.9 (6.2) | 9.3 (5.7) | 9.8 (7.2) | 0.76 |

**Legend:** ABI – ankle brachial index, BMI – body mass index, HTN – hypertension, DM – diabetes mellitus, CAD – coronary artery disease, LURN 10-SF Lower Urinary Tract Research Network, AUASI – American Urologic Association Symptom Index

Among women who completed the LUTS questionnaire compared to women with completed LUTS questionnaires and ABI measures (sTable), only one difference was noted with higher rates of cardio kidney metabolic syndrome among women with ABI measures, 16% vs 28%, p=0.003, respectively.

Among the ABI classification groups, 74% (142/192) had a normal ABI, 20% (38/192) had borderline abnormal ABI and 6% (12/192) had abnormal ABI. Associations between ABI classification group and most sociodemographic, obstetric and gynecologic, and behavioral and medical comorbidity variables were not statistically significant (Table 1). Women with diabetes mellitus were more likely to have an abnormal ABI (p=0.03).

Table 1 shows that the mean LUTS-SI 10 score was 7.2 (SD 5.0), the mean AUASI score was 9.0 (SD 6.2) and the mean BH score was 68.0 (SD 20.3). Differences were most pronounced for BH, with a 10-point difference in mean scores between women in the normal and abnormal ABI classification groups, and for the LURN-SI 10, with a 4-point difference in mean scores.

Table 2 shows results from regressions of each BH and LUTS variable on ABI classification groups, adjusting for covariates in a staged fashion. Women with abnormal versus normal ABI scores had significantly higher LURN-SI 10 scores across all models: a 4.5 point (1.6, 7.4) higher score with adjustment for sociodemographics, a 3.5 (0.4, 6.7) higher score with further adjustment for obstetric and gynecologic factors, and a 4.1 (0.7,7.5) higher score with further adjustment for behavioral and medical comorbidities. Borderline abnormal ABI category was not associated with LURN-SI 10 scores in any model. There was no significant association between ABI classification group and AUASI or BH scores (Table 2).

**TABLE 2.** Associations of BH and LUTS scores among Women with Peripheral Arterial Disease (PAD) by ABI Classification Groups Adjusted for Covariates in Separate Models, n=192.

| Regression normal (reference group) vs borderline vs abnormal | Model 1* | Model 2** | Model 3*** |
| --- | --- | --- | --- |
| Women with PAD - BH |  |  |  |
| Borderline ABI | -1.5 (-8.8, 5.8) | -2.9 (-10.0, 4.5) | -2.6 (-10.0, 5.3) |
| Abnormal ABI | -11 (-23.0, 1.1) | -8.6 (-22.0, 4.4) | -9.6 (-24.0, 4.9) |
| Women with PAD – LURN |  |  |  |
| Borderline ABI | -0.01 (-1.8, 1.8) | 0.2 (-1.6, 2.0) | 0.1 (-1.7, 2.0) |
| Abnormal ABI | 4.5 (1.6, 7.4) | 3.5 (0.4, 6.7) | 4.1 (0.7, 7.5) |
| Women with PAD - AUASI |  |  |  |
| Borderline | 0.4 (-1.9, 2.7) | 0.9 (-1.5, 3.3) | 0.8 (-1.7, 3.2) |
| Abnormal | 1.1 (-2.7, 5.0) | 1.4 (-2.7, 5.5) | 1.4 (-3.1, 5.7) |

\*Model 1 – Sociodemographics
\*\*Model 2 – Obstetric/Gynecologic, including parity, hysterectomy, menopausal status
\*\*\*Model 3 – Behavioral and Medical Comorbidities, including HTN, CAD, DM, Obesity

## DISCUSSION

In this sample of midlife women from CARDIA with an average age of 63.6 years and similar rates of LUTS and BH compared to contemporary estimates,^1,35^ there was a significant relationship between greater LUTS severity and PAD, compared to women with normal ABI. However, LUTS measured with the AUASI and global bladder health did not differ. The relationship with LUTS measured by the LURN-10 and PAD represents preliminary evidence that atherosclerosis or vascular aging at the macrovascular level may be a marker for similar dysfunction to the bladder at the microvascular level. It is possible that both conditions may share similar risk factors. Uniquely, our epidemiologic findings expand vascular geroscience principles from animal models of pelvic ischemia to women with PAD and increased LUTS, as a potential interconnected process.

### Similarities and differences to other studies, including animal studies

Cardiovascular disease (CVD) and CVD risk factors have been associated with an increased prevalence of LUTS across the population.^8,36,37^ Yet it is unknown whether this association is through shared risk factors or shared mechanisms.^9,24,25,27,38–46^ This study gets a step closer to understanding the relationship of LUTS to CVD, specifically with ABI, an accepted measure of PAD and an inexpensive and non-invasive tool. Traditional CVD risk factors, such as metabolic syndrome, nicotine use disorder, hypertension, diabetes mellitus, obesity and hyperlipidemia are associated with increased LUTS prevalence and severity.^9,27,38–46^ In other studies, LUTS has been associated with increased carotid intima-medial thickness and carotid-femoral pulse wave, measures of atherosclerosis, and decreased perfusion to the bladder neck compared to not having LUTS.^21–25^ Further, in animal studies, induced atherosclerosis of the iliac arteries leads to decreased bladder perfusion and detrusor overactivity, a hallmark of OAB, which is a component of LUTS.^13–20^ Very few studies have evaluated PAD, as measured by ABI scores, and LUTS.

Measuring LUTS with contemporary measures, such as LURN-SI 10 questionnaire, may be a more sensitive measure for in women given that it includes assessments of urinary incontinence and bladder pain, whereas the AUASI does not include questions on urinary incontinence or bladder pain.^47^ Also, the BH questionnaire only assesses perceptions of bladder health as opposed to LUTS symptom severity. The BH questionnaire may not be sensitive enough to measure specific decreases in health, like PAD, unlike LUTS measures.

Despite our findings across the LUTS measures, we did have comparable rates of BH and LUTS in our study that were similar to contemporary estimates when using similar questionnaires.^35^ In the RISE FOR HEALTH study, the mean global BH score was estimated to be 72 (IQR 56,84) among women with a mean age of 50 years, with an observed −0.2 lower score per decade of life.^35^ This is comparable to a mean global bladder BH score of 68 among CARDIA women with a mean age of 63 years who elected to have a clinical evaluation after completing LUTS measures. Also in the RISE Study, the mean LURN-SI 10 score was 6.1 (±5.1) among women aged 55-64. Whereas, women in CARDIA electing to come in for a clinical examination reported a mean score of 7.2 (±5.0).

### Strengths and Limitations

The strengths of this study are the high prevalence of participation among women with completed LUTS questionnaires; including the clinical examination and completed the ABI procedure. Similarly, the women were 47% Black, allowing for improved generalization to the population of the United States. However, the design of CARDIA did not include Asian or Hispanic adults. As is common in a community-based cohort studies, this cohort of women may be slightly healthier than the overall population in the United States. However, women who elected to come into the clinical examination did not differ in terms of LUTS or other health concerns, with the exception of metabolic syndrome. The prevalence of PAD in this sample of CARDIA women was 6%, with an additional 20% having borderline abnormal ABI scores. This is comparable to other similarly aged cohorts.^28,48^ However, the relatively small number of women in the abnormal ABI category limited power to detect significant differences in BH and LUTS scores across ABI classification groups. This study only focused on women so is not generalizable to men. As there are associations of CVD events and LUTS severity in men, as well as PAD differences by sex,^8,9,48^ this study aimed to begin exploring the association of PAD and LUTS severity in women. Further research can extend such analyses to men.

## CONCLUSION

These results represent preliminary evidence of a possible association between PAD and LUTS among women that supports vascular geroscience principles.^12^ It is currently unclear if the association could be due to shared mechanisms or shared risk factors. If LUTS represents a peripheral manifestation of CVD, similar to PAD, then preventive strategies for PAD may also decrease the prevalence and/or the natural history of LUTS over time. And, identification of subclinical vascular aging via non-invasive tools could act as an early marker for mitigating LUTS and maintain bladder health over the lifespan.^49^ Further exploration to confirm an association and the potential mechanisms could lead to a better understanding of how to prevent the development or progression of LUTS as well as provide insights for therapeutic development.

## Data Availability

All data produced in the present study are available upon reasonable request to the authors.

https://biolincc.nhlbi.nih.gov/studies/cardia/

## Author Contributions

**Conceptualization:** ADM, CEL, SSB

**Data Curation:** ADM, KA, CEL, SSB, CSF

**Formal Analysis:** SD, AA, GH

**Funding Acquisition:** ADM, SSB

**Investigation:** All authors

**Methodology:** ADM, WH, KA, CEL, SSB, CSF

**Project Administration:** ADM, KA, CSF, CEL, SSB

**Supervision:** ADM, CEL, GH, SSB

**Writing – Original Draft:** all authors

**Writing – Review & Editing:** all authors

